# Depression Remission and Response Rates and Anxiety Response as a Predictor of Depression Response in a Community ECT Clinic

**DOI:** 10.1101/2021.08.25.21262436

**Authors:** Sara B. VanBronkhorst, Evonne M. Edwards, Ali A. Saleem, Darci L. Evans, Eric D. Achtyes, Louis Nykamp, William J. Sanders

## Abstract

**Objective:** To assess depression response and remission rates with electroconvulsive therapy (ECT) in a community clinic and to identify factors predicting success in treatment.

**Methods:** We identified 35 patients by a retrospective chart review with a diagnosis of major depressive disorder or depressive disorder not otherwise specified (according to the Diagnostic and Statistical Manual of Mental Disorders IV-TR) who were treated with an acute series of ECT at the Pine Rest ECT Clinic from March, 2014 to March, 2015. Clinical variables, demographics, depression response rates (based on Patient Health Questionnaire-9; PHQ-9), and anxiety response rates (based on Generalized Anxiety Disorder 7-item) were analyzed.

**Results:** Depression response (defined as ≥ 50% reduction in PHQ-9 score) and remission rates (defined as final PHQ-9 score < 5) were 54.3% and 31.4%, respectively. This was a highly treatment resistant sample, with an average of 5.3 antidepressant failures prior to initiating ECT. Logistic regression analysis found that depression response rates were predicted by an improvement in anxiety symptoms (odds ratio 1.41; 95% confidence interval, 1.11, 1.78). Additionally, patients with initial severe anxiety scores were less likely than other patients to exhibit a response in depression (*p* = .027).

**Conclusion:** Almost half of this sample of patients with treatment resistant depression did not respond to ECT in this community-based clinic. Patients who experienced a response in anxiety symptoms were more likely to experience a depression response while those with severe anxiety were less likely to respond.

## Introduction

Electroconvulsive therapy (ECT) has been shown to be more effective and to provide a more rapid response in the treatment of depression than both placebo and medications.^1-3^ Despite its robust clinical effects, ECT remains underutilized in patients with depression.^4-6^ Fewer hospitals, particularly those treating largely publicly insured patients, conduct ECT, which has contributed to a decreased utilization of ECT.^7^ Due to a number of additional factors negatively associated with ECT use, including stigma, financial constraints, insurance barriers, legal and political policies, cognitive side effects, and transportation,^8,9^ ECT is often reserved for patients who have failed multiple medication trials (patients with treatment resistant depression).^10,11^ Treatment resistant depression is associated with poorer quality of life, increased mortality and poor clinical outcomes.^12,13^ Once treatment resistance is established, successful therapy becomes more difficult to achieve.^12^

While numerous studies have found robust depression response and remission rates with ECT, these rates are mostly based on controlled clinical trials, as opposed to trials in community settings.^14^ Patients who present for ECT in community settings often have severe, treatment resistant depression and have a higher number of medical and psychiatric comorbidities than patients included in clinical trials.^15,16^ In contrast to the 70% to 90% remission rates commonly cited for ECT, which are often based on highly selected patient populations in clinical trials, remission rates found in a large prospective naturalistic study of community hospitals were 30.3% to 46.7%.^14^ Similarly, response rates among patients who have not responded to one or more adequate medication trials are also lower—closer to 50% to 60%.^17-20^

Optimizing treatment, and understanding which patients would most likely benefit from ECT, is important given that access to ECT can be challenging, with nearly 9 out of 10 general hospitals in the United States not conducting ECT.^7^ Administration of ECT in community clinics may vary in factors such as number of treatments, electrode placement, and length of seizure. However, overall, few characteristics have been identified as clinically useful predictors for successful ECT treatment.^18^ This study sought to examine depression response and remission rates, as well as clinical predictors of response to ECT, in a community setting.

## Methods

### Study Participants

Pine Rest Christian Mental Health Services (PRCMHS), located in Grand Rapids, Michigan, is a large, not-for-profit, free-standing psychiatric system. PRCMHS offers a spectrum of comprehensive psychiatric services ranging from inpatient and partial hospitalization to a network of outpatient clinics in the surrounding community. Patients are referred to the Pine Rest ECT Clinic from regional inpatient and outpatient settings. We performed a retrospective chart review to examine depression response and remission rates and to look at clinical factors associated with depression response. Medical records for patients treated with an acute series of ECT at Pine Rest ECT Clinic were reviewed. Approval for this study was obtained from Mercy Health Saint Mary’s Institutional Review Board.

Eligible participants were patients age 18 and older who were treated with an acute series of ECT from March 1, 2014 to March 9, 2015. Inclusion criteria were a primary diagnosis of major depressive disorder or depressive disorder not otherwise specified (NOS) according to the Diagnostic and Statistical Manual of Mental Disorders (DSM) IV-TR.^21^ If a patient had received ECT treatments in the past, there must have been at least six months between the current treatment course and previous treatments. Patients with an initial score of less than 20 on the nine item Patient Health Questionnaire (PHQ-9)^22^ and patients with a DSM IV-TR diagnosis of bipolar disorder, schizophrenia, schizoaffective disorder, or dementia were excluded. Thirty-five patients met criteria for inclusion.

### ECT Treatment Procedure

The acute series of ECT treatment involved treatments three times per week, typically on a Monday, Wednesday, and Friday schedule. A complete acute series was defined as completion of 12 treatments. Patients were started with bilateral or right unilateral electrode placement, based on the recommendation of the primary treating psychiatrist or the psychiatrist providing the second opinion for ECT. Based on clinical discretion, some patients were switched from bilateral to right unilateral electrode placement, others switched from right unilateral to bilateral electrode placement, and others did not switch electrode placement during the course of treatment. Patients could receive ECT treatment while psychiatrically hospitalized or on an outpatient basis.

All patients were treated using the Thymatron System IV machine, manufactured by Somatics, LLC, with a pulse width of 0.50 ms and a current of 0.90 A. Methohexital was the medication of choice for induction of anesthesia, but etomidate was used if there was difficulty achieving seizure, if the patient was taking an antiepileptic medication or benzodiazepine, or if the patient had received ECT previously with etomidate. Succinylcholine was used to induce paralysis. Charge dose was determined by the psychiatrist providing ECT treatment. In general, a therapeutic dose was considered approximately 1-1/2 to 2-1/2 times the seizure threshold when giving bilateral treatments and approximately 5 to 7 times the seizure threshold when giving right unilateral treatments.

### Data Collection and Outcome Measures

Study data including sex, age, diagnosis, electrode placement, number of treatments, number of antidepressant medication trials, and scores on assessment tools, were obtained by review of the medical record for the first 12 treatments received during the acute series. Patients completed the PHQ-9 and the Generalized Anxiety Disorder 7-item scale (GAD-7)^23^ prior to each ECT treatment. The Hamilton Depression Rating Scale (HAMD-17)^24^ and the Montreal Cognitive Assessment (MoCA)^25^ were administered before the first, sixth, and last ECT treatments. Response for depression was defined as a 50% or greater reduction of the PHQ-9 score. A secondary measure of response for depression was a 50% or greater reduction in the HAMD-17 score. Response for anxiety was defined as a 50% or greater reduction in GAD-7 scores. Remission for depression was defined as PHQ-9 score < 5, with a secondary measure for remission being a score ≤ 7 on the HAMD-17. Change scores were calculated for PHQ-9 and GAD-7 scores by subtracting final scores from initial scores for each patient at the conclusion of the acute series of ECT.

### Statistical Analysis

Descriptive statistics were analyzed for demographic and treatment variables. Based on previously reported response rates of 50% to 60% for patients who had not responded to one or more adequate medication trials,^17-20^ chi-squared goodness-of-fit tests were used to compare response rates in this study to a 55% response rate. Chi-squared goodness-of-fit tests were also utilized to assess remission rates and used a comparison remission rate of 38.5%, the midpoint of the remission rates range found in a prior study of ECT in community hospital settings.^14^ Chi-squared tests of independence were used to compare response rates between patients differing in demographic and treatment variables, including initial electrode placement, initial treatment location, age, MoCA scores, and initial anxiety scores (above or below screening score). Additional logistic regression analyses were utilized to determine predictors of PHQ-9 response. Patients missing the first or second HAMD-17 score were excluded from analyses. When final scores for HAMD-17 or PHQ-9 were missing, last observation carried forward was used for analyses.

## Results

### Patient Demographics and Clinical Features

A total of 35 patients met inclusion criteria (22 women and 13 men, mean age 52.9; *SD* = 12.9). The mean number of treatments was 9.3 (*SD* = 3.0). See Table 1 for demographics and descriptive characteristics with depression response rates. An exhaustive previous medication trial history, including duration and dose of medication use, was not elicited by psychiatric providers in a standardized manner; however, using available data in psychiatric assessments, the mean number of antidepressant medication trials was found to be 5.3 (*SD* = 2.4). Twenty-four patients (68.6%) started treatment while on an inpatient psychiatric unit, and 22 (62.9%) started with bilateral electrode placement. Thirteen patients completed at least 12 treatments, and of these, 6 responded and 2 remitted. Of the 21 patients who did not complete 12 treatments, 13 responded and 9 remitted. Eight patients dropped out of treatment without responding before completing 12 treatments. See Figure 1 for a trajectory of mean PHQ-9 score by depression improvement outcomes.

**Table 1.**
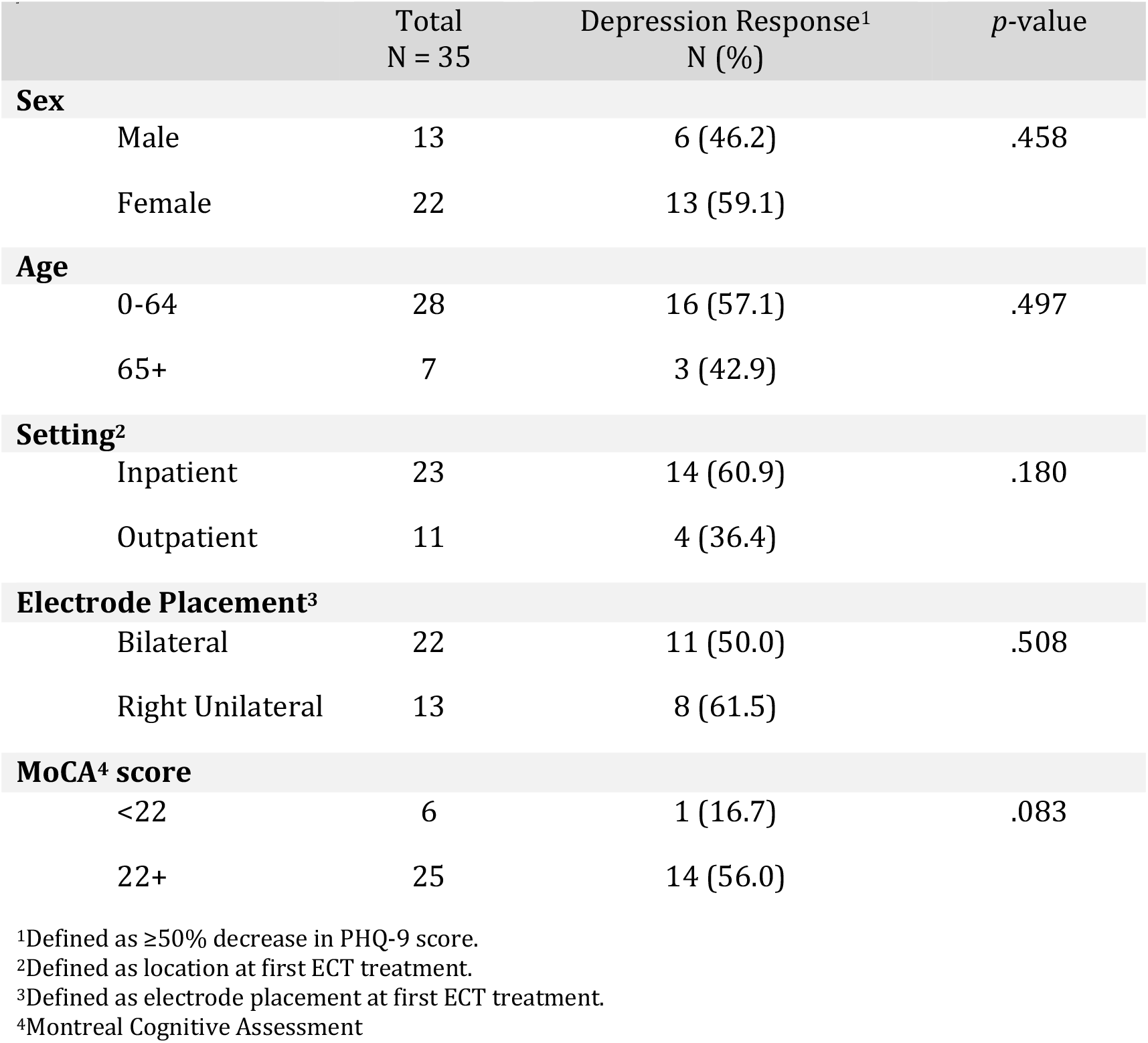
Depression Response Rates by Demographics and Descriptive Characteristics in Community ECT Clinic

|  | Total<br>N = 35 | Depression Response<sup>1</sup><br>N (%) | *p*-value |
| --- | --- | --- | --- |
| Sex |  |  |  |
| Male | 13 | 6 (46.2) | .458 |
| Female | 22 | 13 (59.1) |  |
| Age |  |  |  |
| 0-64 | 28 | 16 (57.1) | .497 |
| 65+ | 7 | 3 (42.9) |  |
| Setting<sup>2</sup> |  |  |  |
| Inpatient | 23 | 14 (60.9) | .180 |
| Outpatient | 11 | 4 (36.4) |  |
| Electrode Placement<sup>3</sup> |  |  |  |
| Bilateral | 22 | 11 (50.0) | .508 |
| Right Unilateral | 13 | 8 (61.5) |  |
| MoCA<sup>4</sup> score |  |  |  |
| <22 | 6 | 1 (16.7) | .083 |
| 22+ | 25 | 14 (56.0) |  |

<sup>1</sup>Defined as ≥50% decrease in PHQ-9 score.
<sup>2</sup>Defined as location at first ECT treatment.
<sup>3</sup>Defined as electrode placement at first ECT treatment.
<sup>4</sup>Montreal Cognitive Assessment

**Figure 1.**
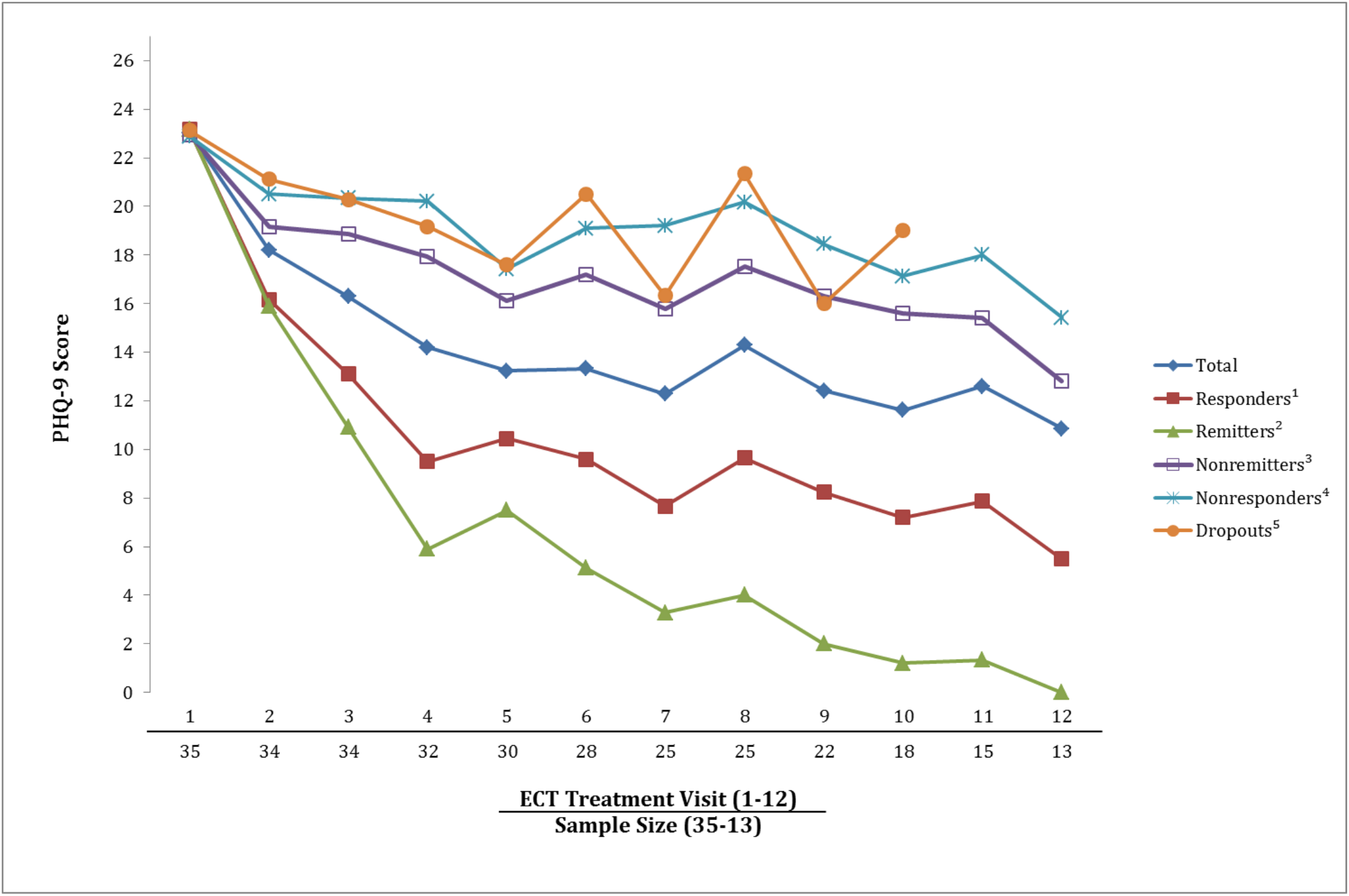
Trajectory of Mean Patient Health Questionnaire (PHQ-9) Scores, by Outcome Group, in Community ECT Clinic ^1^Defined as ≥50% decrease in PHQ-9 score. ^2^Defined as final PHQ-9 score <5. ^3^Defined as completing 12 treatments without ≥50% decrease in PHQ-9 score. ^4^Defined as completing 12 treatments with final PHQ-9 score ≥5. ^5^Defined as completing <12 treatments without ≥50% decrease in PHQ-9 score.

Electrode placement was based on clinical judgement by the treating psychiatrist and a total of 8 patients switched electrode placement during the course of treatment. Of the 22 patients who started with bilateral treatment, 3 patients switched to right unilateral treatment; of the 13 patients who started with right unilateral, 5 switched to bilateral. Among patients who started with bilateral treatment, 11 (50.0%) had a response in their depressive symptoms, compared to 8 of the 13 patients who started with right unilateral (61.5%); however, this difference was not statistically significant, X ^*2*^ (1, *N =* 35) = .44, *p* = .51. Eight of the 23 patients who started on an inpatient psychiatric unit switched to outpatient. Response rates on the PHQ-9 for those patients who started ECT while inpatient (67.6%) did not significantly differ from response rates for patients who started ECT while outpatient (32.4%), X ^*2*^ (1, *N =* 34) = 1.79, *p* = .18. Similarly, there was no difference found in PHQ-9 response rates when comparing those age 65 and older (42.9%) with those younger than 65 (57.1%), *X* ^*2*^ (1, *N* = 35) = .461, *p* = .497.

Although patients with a diagnosis of dementia were excluded from the study, 6 of the 31 patients who completed a MoCA (19.4%), had a score of < 22 prior to ECT, which is a score consistent with a likely diagnosis of neurocognitive disorder.^26^ While only 16.7% of patients with a MoCA score of < 22 responded on the PHQ-9, 56.0% of those with a score of ≥ 22 responded; this difference approached statistical significance, X ^*2*^ (1, *N =* 31) = 3.0, *p* = .083. Similarly, those with a score of < 22 were found to be less likely to respond on the HAMD-17 (0%) than those with a score of 22 or greater (57.1%), X ^*2*^ (1, *N =* 25) = 4.40, *p* = .036.

### Response and remission rates

Overall, 19 patients (54.3%) had depression response on the PHQ-9, which was the primary response measure. This response rate did not differ from previously published response rates for patients who had not responded to one or more adequate medication trials (55%), with X^*2*^ (1, *N =* 35) = 0.007, *p* = .932. A similar response rate of 57.1% was seen on the HAMD-17, which did not differ from a 55% response rate, X ^*2*^ (1, *N =* 26) = 0.822, *p* = .365. A total of 11 patients (31.4%) met remission criteria as measured by PHQ-9, and 23.1% of patients remitted as measured by the HAMD-17. These remission rates based on the PHQ-9 and HAMD-17 did not significantly differ from 38.5%, the comparison remission rate estimated from previously published studies, with X ^*2*^ (1, *N =* 35) = 0.739, *p* = .390, and *X* ^*2*^ (1, *N =* 26) = 2.612, *p* = .106, respectively.

Nineteen patients (54.3%) had response of anxiety symptoms, as measured by 50% reduction in GAD-7 score. Among patients who exhibited a response on the PHQ-9, 84.2% also exhibited a response on the GAD-7. Logistic regression analysis found that both PHQ-9 and HAMD-17 response rates were significantly predicted by improvements in GAD-7, with odds ratios (ORs) of 1.407, (CI 95% 1.113, 1.779), *p* = .004, and OR of 1.281, (CI 95% 1.033, 1.588), *p* = .024, respectively. Initial GAD-7 scores did not predict PHQ-9 response rates, OR = 0.950, (CI 95% 0.812, 1.112), *p* = .523. Additionally, correlations between initial GAD-7 and PHQ-9 scores, *r*(33) = .299, *p* = .081, were not statistically significant and were of a lesser magnitude than the correlation between GAD-7 change scores and PHQ-9 change scores, *r*(33) = .744, *p* < .001, indicating that amount and direction of change in one measure was positively associated with amount and direction of change in the other measure. Initial PHQ-9 scores did not predict PHQ-9 response rates, OR = 1.062, (CI 95% 0.780, 1.446), *p* = .701, suggesting that among this group of patients with severe depression, initial depression severity was not predictive of response to treatment. While those patients who screened positive for an anxiety disorder (initial GAD-7 ≥ 10) did not differ in depression response rates from those screening negative for an anxiety disorder *X* ^*2*^ (1, *N* = 35) = 0.203, *p* = .653, patients with initial severe anxiety scores (GAD-7 ≥ 15) were less likely than other patients to exhibit a response in depression as measured by the PHQ-9; *X* ^*2*^ (1, *N* = 35) = 4.900, *p* = .027, (41.7% vs. 81.8%).

## Discussion

The response and remission rates for depression with ECT treatment in this sample (54.3% and 31.4%, respectively) were comparable to previously reported rates among patients who had not responded to one or more adequate medication trials (50% to 60%),^17-20^ and the results were also similar to findings from a large prospective study in community hospitals (30.3% to 46.7%).^14^ Although there was not a standardized, thorough assessment of previous medication trials completed on every patient, available data in admission notes found the average number of antidepressants trials was 5.3, suggesting that the patients in this study had significant medication resistance. Not only is resistance to medication trials a predictor of poor response to ECT treatment, but the degree of resistance to previous treatments has been found to be the strongest predictor of nonresponse to ECT.^27^

While success rates for depression treatment were similar to results from comparable studies, these rates are lower than the robust success rates of 70% to 90% commonly touted for ECT treatment of depression. This underscores the clinical challenge encountered with treatment resistant depression and with patients presenting for treatment in community ECT clinics. Several explanations have been suggested for the discrepancy between outcomes in prospective research trials compared to community treatments with ECT, with earlier discontinuation of treatments in community settings having been proposed as a likely contributor.^14^ In this study, 8 of the 35 patients dropped out of treatment without responding. Reasons for early patient discontinuation should be further evaluated as this may contribute to lower than expected depression response rates. Another possible explanatory factor is that patients undergoing ECT in clinical trials may be more likely to be taking prescribed medications during the course of ECT than non-research patients. Similarly, patients who self-select for participation in a clinical trial may be more likely to be adherent to treatment recommendations than those in unselected community samples.^28,29^ It should be noted that while the results are not as robust as would be hoped for, the ECT response rates were still higher than those found for additional medication trials in patients with treatment resistant depression,^13^ suggesting ECT is likely underutilized and not used early enough in a patient’s treatment course in patients with treatment resistant depression. This study highlights the need to find ways to maximize ECT benefit and identify factors which predict ECT success.

In this study, an association between improvements in anxiety and depression during the course of ECT treatment was found. Anxiety is often comorbid with depression, with the United States National Comorbidity Survey Replication study finding the prevalence of 12-month comorbidity of an anxiety disorder among those with major depressive disorder to be 53.3% to 61.7%.^30^ Additionally, in patients with depression, comorbid anxiety has been found to adversely affect outcomes not only with medication treatment,^31,32^ but also electroconvulsive therapy.^33,34^ As of yet, limited data regarding anxiety symptom response with ECT exists,^35-37^ and given its prevalence in patients with depression, this warrants further investigation. We found that patients who experienced a response in depressive symptoms were also more likely to demonstrate a response in anxiety symptoms. This relationship may be partially accounted for by known correlations between self-reported anxiety and depression. However, correlations between initial GAD-7 and PHQ-9 scores were not statistically significant and were of a lesser magnitude than those between GAD-7 change scores and PHQ-9 scores. This suggests a relationship between improvements in anxiety and depression with ECT treatment. If this is a consistent relationship, questions arise regarding its etiology. Is ECT itself treating anxiety symptoms in those whose depression responds to ECT treatment? Or perhaps the improvement in anxiety symptoms contributes to the alleviation of depressive symptoms. If so, would aggressively treating anxiety during ECT treatment improve depression outcomes?

Patients with high anxiety scores were less likely to experience a response in depressive symptoms than were other patients, with approximately half as many patients with high anxiety experiencing response than those without high anxiety. Of note, these findings were based on anxiety evaluated at first ECT treatment and thus did not differentiate between anxiety related to starting ECT and anxiety unrelated to ECT treatment. However, assessing anxiety level prior to first ECT treatment may be informative in predicting which patients are more likely to have depression that is responsive to ECT treatment. While severity of depression (assessed by initial PHQ-9 scores) did not predict depression response rates, this may be due, in part, to a restricted range of initial PHQ-9 scores due to excluding patients with initial PHQ-9 scores below 20. While previous studies have found demographic factors^38^ and electrode placement^39^ to predict depression response, this study did not find these factors to be predictive of depression response. This may be due to a small sample size that limited the power to detect effects of electrode placement, age, and sex.

To help address some of these questions, future research recommendations include evaluating depression response rates in a prospective ECT trial involving adjunctive treatment of anxiety. Evaluating anxiety scores prior to referral to ECT may help differentiate baseline anxiety from anxiety related to ECT. There is a paucity of research evaluating ECT for treatment of anxiety in general, and further evaluating this would be of clinical benefit.

There are several limitations to this study. The small sample size limited the power in analyses. Additionally, only 13 of the 35 patients completed 12 sessions and data regarding reason for early discontinuation was missing. Finally, there was a lack of standardization in the provision of the ECT treatment and psychiatrists administering treatment may not have titrated treatments in the same way or made recommendations about right unilateral versus bilateral electrode placement consistently. Due to the limited sample size, we were unable to evaluate these potential effects by provider or treatment titration approach.

## Conclusions

Approximately half of the patients in this community sample of patients with highly treatment resistant depression responded with ECT treatment, and one third achieved remission. These results, although comparable to success rates of patients in community settings, are lower than what are commonly cited with ECT in clinical trials. Anxiety response rates were similar to depression response rates and improvement in anxiety symptoms was associated with response in depressive symptoms. Fewer patients with severe anxiety achieved depression response than those without severe anxiety. Further work is needed to understand the reasons for lower response and remission rates in community settings as well as the impact of anxiety on treatment response with ECT.

## Data Availability

Data can be made available on request

## References

1. Fink M. What was learned: studies by the consortium for research in ECT (CORE) 1997-2011. Acta Psychiatrica Scandinavica. 2014;129(6):417–426.

2. Kho KH, van Vreeswijk MF, Simpson S, Zwinderman AH. A meta-analysis of electroconvulsive therapy efficacy in depression. The Journal of ECT. 2003;19(3):139–147.

3. Carney S, Cowen P, Dearness K, Eastaugh J, UK ECT Review Group. Efficacy and safety of electroconvulsive therapy in depressive disorders: a systematic review and meta-analysis. The Lancet. 2003;361:799–808

4. Wilkinson ST, Agbese E, Leslie DL, Rosenheck RA. Identifying recipients of electroconvulsive therapy: data from privately insured Americans. Psychiatric Services. 2018. doi: 10.1176/appi.ps.201700364.

5. Sackeim H. Modern electroconvulsive therapy: vastly improved yet greatly underused. JAMA Psychiatry. 2017;74(8):779–780.

6. Taylor S. Electroconvulsive therapy: a review of history patient selection, technique, and medication management. Southern Medical Journal. 2007;100(5):494–498.

7. Case BG, Bertollo DN, Laska EM, et al. Declining use of electroconvulsive therapy in United States general hospitals. Biological Psychiatry. 2013;73(2):119–126.

8. Livingston R, Anandan S, Moukaddam N. Electroconvulsive therapy, transcranial magnetic stimulation, and deep brain stimulation in treatment-resistant depression. Psychiatric Annals. 2016;46(4):240–246.

9. Philpot M, Treloar A, Gormley N, Gustafson L. Barriers to the use of electroconvulsive therapy in the elderly: a European survey. European Psychiatry. 2002;17(1):41–45.

10. Heijnen W, Birkenhager T, Wierdsma A, van den Broek W. Antidepressant pharmacotherapy failure and response to subsequent electroconvulsive therapy: a meta-analysis. Journal of Clinical Psychopharmacology. 2010;30(5):616–619.

11. Alpert JE. Improving depression outcome: new concepts, strategies and technologies. Journal of Psychopharmacology. 2006;20(3):3–4.

12. Fekadu A, Wooderson S, Markopoulo K, Donaldson C, Papadopoulos A, Cleare A. What happens to patients with treatment-resistant depression? a systematic review of medium to long term outcome studies. Journal of Affective Disorders. 2009;116(1-2):4–11.

13. Sinyor M, Schaffer A, Levitt A. The sequenced treatment alternatives to relieve depression (STAR* D) trial: a review. The Canadian Journal of Psychiatry. 2010;55(3): 126–135.

14. Prudic J, Olfson M, Marcus SC, Fuller RB, Sackeim HA. Effectiveness of electroconvulsive therapy in community settings. Biological Psychiatry. 2004;55(3):301–312.

15. Abrams R. Electroconvulsive Therapy. 4th ed. New York, NY: Oxford University Press; 2002.

16. American Psychiatric Association. The practice of electroconvulsive therapy: recommendations for treatment, training, and privileging: a task force report of the American Psychiatric Association. 2nd ed. Washington, D.C: American Psychiatric Association; 2001.

17. Sackeim HA, Prudic J, Devanand DP, et al. A prospective, randomized, double-blind comparison of bilateral and right unilateral electroconvulsive therapy at different stimulus intensities. Archives of General Psychiatry. 2000;57(5):425–434.

18. Haq AU, Sitzmann AF, Goldman ML, Maixner DF, Mickey BJ. Response of depression to electroconvulsive therapy: a meta-analysis of clinical predictors. The Journal of Clinical Psychiatry. 2015;76(10):1374–1384.

19. Prudic J, Haskett RF, Mulsant B, et al. Resistance to antidepressant medications and short-term clinical response to ECT. American Journal of Psychiatry. 1996;153(8):985–992.

20. Devanand DP, Sackeim HA, Prudic J. Electroconvulsive therapy in the treatment-resistant patient. Psychiatric Clinics of North America. 1991;14(4):905–23.

21. American Psychiatric Association. Diagnostic and statistical manual of mental disorders: DSM-IV-TR. 4th ed., text revision. Washington, DC: American Psychiatric Association; 2000.

22. Kocalevent RD, Hinz A, Brähler E. Standardization of the depression screener patient health questionnaire (PHQ-9) in the general population. General Hospital Psychiatry. 2013;35(5):551–555.

23. Spitzer RL, Kroenke K, Williams JBW, Löwe B. A brief measure for assessing generalized anxiety disorder: the GAD-7. Archives of Internal Medicine. 2006;166(10):1092–1097.

24. Leentjens AF, Verhey FR, Lousberg R, Spitsbergen H, Wilmink FW. The validity of the Hamilton and Montgomery-Asberg depression rating scales as screening and diagnostic tools for depression in Parkinson’s disease. International Journal of Geriatric Psychiatry. 2000;15(7):644–649.

25. Freitas S, Simões MR, Alves L, Santana I. Montreal cognitive assessment: validation study for mild cognitive impairment and Alzheimer disease. Alzheimer Disease and Associated Disorders. 2013;27(1):37–43.

26. Borland E, Nägga K, Nilsson PM, Minthon L, Nilsson ED, Plamqvist S. The Montreal cognitive assessment: normative data from a large Swedish population-based cohort. Journal of Alzheimer’s Disease. 2017;59(3):893–901.

27. Milev RV, Giacobbe P, Kennedy SH, et al. Canadian network for mood and anxiety treatments (CANMAT) 2016 clinical guidelines for the management of adults with major depressive disorder: section 4. neurostimulation treatments. The Canadian Journal of Psychiatry. 2016;61(9):561–575.

28. Ten Have TR, Normand ST, Marcus SM, Brown CH, Lavori P, Duan N. Intent-to-treat vs. non-intent-to-treat analyses under treatment non-adherence in mental health randomized trials. Psychiatric Annals. 2008;38(12):772–783.

29. Schulberg HC, Block MR, Madonia MJ, et al. Treating major depression in primary care practice: eight-month clinical outcomes. Archives of General Psychiatry. 1996;53(10):913–919.

30. Kessler RC, Berglund P, Demler O, et al. The Epidemiology of Major Depressive Disorder: Results From the National Comorbidity Survey Replication (NCS-R). JAMA. 2003;289:3095–3105.

31. Rivas-Vazquez RA, Saffa-Biller D, Ruiz I, Blais MA, Rivas-Vazquez A. Current issues in anxiety and depression: comorbid, mixed, and subthreshold disorders. Professional Psychology: Research and Practice. 2004;35(1):74–83.

32. Pollack MH. Comorbid anxiety and depression. The Journal of Clinical Psychiatry. 2005;66(suppl 8):22–29.

33. Brus O, Cao Y, Gustafsson E, et al. Self-assessed remission rates after electroconvulsive therapy of depressive disorders. European Psychiatry. 2017;45:154–160.

34. Berggren Å, Gustafson L, Höglund P, et al. A long-term follow-up of clinical response and regional cerebral blood flow changes in depressed patients treated with ECT. Journal of Affective Disorders. 2014;167:235–243.

35. Obbels J, Verwijk E, Bouckaert F, Sienaert P. ECT-related anxiety: a systematic review. The Journal of ECT. 2017;33(4):229–236.

36. Ozdemir O, Yillmaz E, Atilla E. Is electroconvulsive therapy (ECT) effective in the treatment of psychosis or anxiety disorders? report of two cases. Journal of Mood Disorders. 2014;4(3):122–125.

37. Kong XM, Xu SX, Sun Y, et al. Electroconvulsive therapy changes the regional resting state function measured by regional homogeneity (ReHo) and amplitude of low frequency fluctuations (ALFF) in elderly major depressive disorder patients: an exploratory study. Psychiatry Research. 2017; 264:13–21.

38. van Diermen L, van den Ameele S, Kamperman A, et al. Prediction of electroconvulsive therapy response and remission in major depression: meta-analysis. The British Journal of Psychiatry. 2018;212(2):71–80.

39. Kellner CH, Tobias KG, Wiegand J. Electrode placement in electroconvulsive therapy (ECT): a review of the literature. The Journal of ECT. 2010;26(3):175–180.

